# Rapidly increasing SARS-CoV-2 seroprevalence and limited clinical disease in three Malian communities: a prospective cohort study

**DOI:** 10.1101/2021.04.26.21256016

**Authors:** Issaka Sagara, John Woodford, Mamady Kone, Mahamadoun Hamady Assadou, Abdoulaye Katile, Oumar Attaher, Amatigue Zeguime, M’Bouye Doucoure, Emily Higbee, Jacquelyn Lane, Justin Doritchamou, Irfan Zaidi, Dominic Esposito, Jennifer Kwan, Kaitlyn Sadtler, Alassane Dicko, Patrick Duffy

## Abstract

**Background:** The extent of SARS-CoV-2 exposure and transmission in Mali and the surrounding region is not well understood, although infection has been confirmed in nearly 14,000 symptomatic individuals and their contacts since the first case in March 2020. We aimed to estimate the cumulative incidence of SARS-CoV-2 in three Malian communities, and understand factors associated with infection.

**Methods:** Between 27 July 2020 and 29 January 2021, we collected blood samples along with demographic, social, medical and self-reported symptoms information from residents aged 6 months and older in three study communities at two study visits. SARS-CoV-2 antibodies were measured using a highly specific two-antigen ELISA optimized for use in Mali. We calculated cumulative adjusted seroprevalence for each site and evaluated factors associated with serostatus at each visit by univariate and multivariate analysis.

**Findings:** Overall, 94.8% (2533/2672) of participants completed both study visits. A total of 50.3% (1343/2672) of participants were male, and 31.3% (837/2672) were aged <10 years, 27.6% (737/2672) were aged 10-17 years, and 41.1% (1098/2572) were aged ≥18 years. The cumulative SARS-CoV-2 exposure rate was 58.5% (95% CI: 47.5 to 69.4). This varied between sites and was 73.4% (95% CI: 59.2 to 87.5) in the urban community of Sotuba, 53.2% (95% CI: 42.8 to 63.6) in the rural town of Bancoumana, and 37.1% (95% CI: 29.6 to 44.5) in the rural village of Donéguébougou. This equates to an infection rate of approximately 1% of the population every three days in the study communities between visits. Increased age and study site were associated with serostatus at both study visits. There was minimal difference in reported symptoms based on serostatus.

**Interpretation:** The true extent of SARS-CoV-2 exposure in Mali is greater than previously reported and now approaches hypothetical herd immunity in urban areas. The epidemiology of the pandemic in the region may be primarily subclinical and within background illness rates. In this setting, ongoing surveillance and augmentation of diagnostics to characterize locally circulating variants will be critical to implement effective mitigation strategies like vaccines.

**Funding:** This project was funded by the Intramural Research Program of the National Institute of Allergy and Infectious Diseases, National Institute of Biomedical Imaging and Bioengineering, and National Cancer Institute.

## INTRODUCTION

Many African nations have seemingly been spared the overwhelming burden of disease seen in other countries during the first waves of the COVID-19 pandemic. This may be attributed to a younger population age structure and other hypothetical but undefined host or virological factors [1, 2].

In Mali, the first cases of COVID-19 were detected in March 2020, and as of 5 April 2021 there have been 10,622 confirmed cases from 241,431 viral detection tests. The true extent of SARS-CoV-2 infection in many African nations is likely to be greater than previously reported. Understanding the extent of infection and burden of disease is critical to allocate limited public health resources, including vaccines. Case numbers may be underestimated due to asymptomatic and paucisymptomatic infections, as well as healthcare access and diagnostic capacity.

Serosurveillance is a convenient and potentially powerful tool to understand the extent of SARS-CoV-2 infection in the community. Despite the large number of serological assays available globally, reporting methods have not been standardized nor have assays routinely been qualified for use in populations under study, hence SARS-CoV-2 seroprevalence information may be inconsistent and have uncertain test predictive characteristics. This is particularly relevant in sub-Saharan Africa, where the high infectious disease burden may affect serology interpretation [3-6] and access to laboratory infrastructure is often limited.

Using commercial point of care tests, community serosurveillance throughout 2020 has identified gradually increasing seroprevalence rates in West African countries, including 0.9% in Togo in April, 25.4% in Nigeria in June, and 25.1% in Côte d’Ivoire in October [7-9]. Similarly, surveys in other parts of the continent using laboratory-based single antigen ELISA have estimated seroprevalence rates of 4.3% in Kenyan blood donors in June 2020, 2.1% in households in Zambia in July, and up to 60% in blood donors in parts of South Africa in January 2021 [10-12]. These results suggest that SARS-CoV-2 is circulating throughout Africa, in some cases potentially at a subclinical level, and that there may be a largely unquantified community reservoir of transmission.

We sought to determine the age-specific cumulative incidence of SARS-CoV-2 infection in longitudinal cohorts at urban and rural sites in Mali, using a two-antigen ELISA previously optimized for serodiagnosis in the local population [6]. In addition, we examined demographic, social and medical factors, including self-reported symptoms history, for associations to serostatus, compared seropositivity to seroconversion episodes to further assess assay performance, and characterized the longitudinal dynamics of the antibody response to each of the target antigens.

## METHODS

### Study design and population

This prospective cohort study was adapted from the WHO population-based age-stratified seroepidemiological investigation protocol for COVID-19 virus infection version 1.1 [13]. We assessed individuals aged 6 months or older from three communities in Mali for anti-SARS-CoV-2 antibodies. The participating communities were Sotuba (urban), Bancoumana (rural town), and Donéguébougou (rural villace) (Supplementary Figure 1). Each of these sites has an existing MRTC/NIH study facility engaged with the local community. Individuals participating in existing malaria studies, and residents of the local community were invited to participate.

Sotuba is a community population approximately 7,000 located on the bank of the Niger River, in the capital city Bamako (total population ∼2.7M). Many clinical trials, as well as epidemiological and entomologic malaria studies, have been conducted in Sotuba. Cumulative malaria exposure in Sotuba is modest compared to highly endemic parts of Mali. The most recent census data available were collected in 2017.

Bancoumana town is located 60 kilometers southwest of Bamako on the main road to Guinea-Conakry and has a population of approximately 10,000 people. The site is situated in the south-Sudanian area of Mali. The climate is hot, with daily temperatures ranging from 19°C to 40°C. Many clinical trials, as well as epidemiological and entomologic malaria studies, have been conducted in Bancoumana. Malaria transmission is highly seasonal and intense during the rainy season from July to December. The most recent census data available were collected in 2018.

Donéguébougou is a village located 30 km north of Bamako and has a population of approximately 2,000 people. For the purpose of malaria vaccine trials and epidemiology studies, facilities have been put in place at Donéguébougou within walking distance to the residents’ homes. There is a high study participation rate per compound in Donéguébougou, thus this site is well suited for community-wide assessments. Malaria transmission is highly seasonal and intense, with the transmission season taking place from June until December. The most recent census data available were collected in 2019.

### Ethics Statement

The study was conducted as a Public Health surveillance activity in collaboration with the Malian Ministry of Health and was approved by the ethics committee of Facultes de Medicine/d’Odonto-Stomatologie et de Pharmacie (2020/114/CE/FMOS/FAPH) and the Malian COVID-19 Scientific Review Committee. Written informed consent or assent was obtained from all participants before enrollment in the study.

### Procedures

Participants were invited to complete two study visits. Visit 1 occurred at enrollment commencing 28 July 2020 and visit 2 occurred commencing 14 December 2020. Demographic characteristics, symptom history, medical comorbidities, and social history were collected from participants. Infants aged 6-12 months were co-enrolled with participating mothers. At each visit, 3.5-10 mL venous blood samples were collected. Data were collected and stored using REDCap. Participants were provided with a re-usable mask supplied by the Ministry of Health and were requested to observe physical distancing. Study staff were required to use personal protective equipment and environmental controls in line with site standard operating procedures.

Sera separated from blood samples collected at each visit were tested for the presence of IgG antibodies to SARS-CoV-2 spike protein and receptor binding domain (RBD) at the MRTC/DEAP Immunology Laboratory using a reference ELISA adapted for optimized performance in the local population [6, 14]. Seropositivity was defined as spike protein and RBD assay absorbance values (optical density, OD) above antigen cutoffs. The estimated sensitivity and specificity of these cutoffs was 73.9% (51.6 to 89.8) and 99.4% (97.7 to 99.9) respectively [6]. To further evaluate the performance of the assay, a subset of study participants with pre-pandemic samples available were evaluated for spike protein and RBD seroconversion. All samples were tested in duplicate, with plate negative controls (pooled pre-pandemic sera) and plate positive controls (monoclonal antibody CR3022) by trained laboratory staff. Study samples with discordant duplicate results (>20%) or results around the assay cutoffs were repeated.

### Covariates

Demographic variables included age, sex, and community of residence. Medical comorbidity was defined as the presence of at least one of the following self-reported conditions: obesity, diabetes, human immunodeficiency virus (HIV) or other immunosuppression, hypertension, heart disease, chronic pulmonary disease, chronic liver disease, chronic hematologic disorder, chronic kidney disease, chronic neurological disease, and malignancy. Participants also reported smoking status, history of Bacillus Calmette-Guérin (BCG) vaccination and recent antimalarial use (<4 weeks). In female participants, pregnancy status was recorded. Social history included employment in a healthcare facility, household member employment in a healthcare facility, household member diagnosed with COVID-19, and household size. Symptom history included systemic symptoms: fever, chills, fatigue, myalgia, and headache; respiratory symptoms: sore throat, cough, rhinorrhea, shortness of breath, wheeze, anosmia/loss of taste, and respiratory symptoms not otherwise specified; and gastrointestinal symptoms: nausea/vomiting, abdominal pain, and diarrhea. Symptom severity was estimated based on self-reported school or work absenteeism, presentation for medical attention, and hospitalization. Infants aged 6 to 12 months were co-enrolled with their mother, and a limited history was collected.

### Statistical analysis

The study sample size was based on pragmatic factors. A provisional target of 500-1000 participants per site was requested to allow for age stratification [13]. Seroprevalence and 95% confidence intervals for each site were calculated for each visit. Seroprevalence estimates were adjusted using two methods. Firstly, results for each site were stratified by age group (<10 years, 10-17 years, ≥18 years) and weighted for community age structure and size using available census data. Secondly, results were adjusted for test sensitivity and specificity [15]. The cumulative adjusted SARS-CoV-2 exposure prevalence at visit 2 was estimated by including seropositive cases from visit 1 that had seroreverted at visit 2 before applying adjustments. The daily rate of infection was estimated by calculating the adjusted incidence of new cases between visit 1 and visit 2 and dividing this number by the median number of days between visits. Chi squared tests were used to test for differences between site seroprevalence estimates.

### Exploratory analyses

To assess assay performance, seroconversion episodes were calculated for participants with stored pre-pandemic samples available for assessment. Seroconversion was defined as a fourfold increase in absorbance value at visit 1 compared to a paired pre-pandemic sample for SARS-CoV-2 spike protein and RBD. Very low absorbance values were replaced with the assay limit of blank [16] to improve the accuracy of seroconversion estimates. To confirm the performance of the assay cutoffs, concordance between crude seropositivity and seroconversion was assessed by Cohen’s kappa.

The effect of selected co-variates on serostatus at visit 1 and visit 2 were modeled by multiple logistic regression. Site, age, sex, and self-reported symptoms (by category) were included *a priori*. Other co-variates were selected based on univariate analysis of seronegative and seropositive groups at each visit, using a p value threshold of 0.05. Fisher exact tests were used for categorical variables, unpaired two-tailed t-tests were used for continuous variables. Similar co-variates were grouped by category (for example, nausea/vomiting, abdominal pain, and diarrhea were grouped as gastrointestinal symptoms).

In participants confirmed seropositive at visit 1, the proportion of seroreversions at visit 2 was calculated. To assess antibody kinetics, the rate of change of SARS-CoV-2 spike and RBD absorbance value (OD/100 days) was also calculated.

In a subset of adult participants co-enrolled in clinical trials at the Bancoumana site [17] and the Donéguébougou site [18] clinical trial MedDRA coded adverse events occurring between visit 1 and 7 days before visit 2 were assessed to better understand the clinical presentation of COVID-19. Adverse event rates were compared based on serostatus at visit 2 in all participants seronegative at visit 1 using the Fisher Exact Test.

Statistical analysis was performed with Microsoft Excel and Graphpad Prism 9 software.

## RESULTS

### Study population

A total of 2673 individuals were screened at three study sites, and 2672 were enrolled (Figure 1). At the urban Sotuba site 594 participants including 9 co-enrolled infants paired with their mother were enrolled, at the rural Bancoumana site 965 participants including 6 co-enrolled infants were enrolled, and at the rural Donéguébougou site 1113 participants were enrolled (no co-enrolled infants).

**Figure 1:**
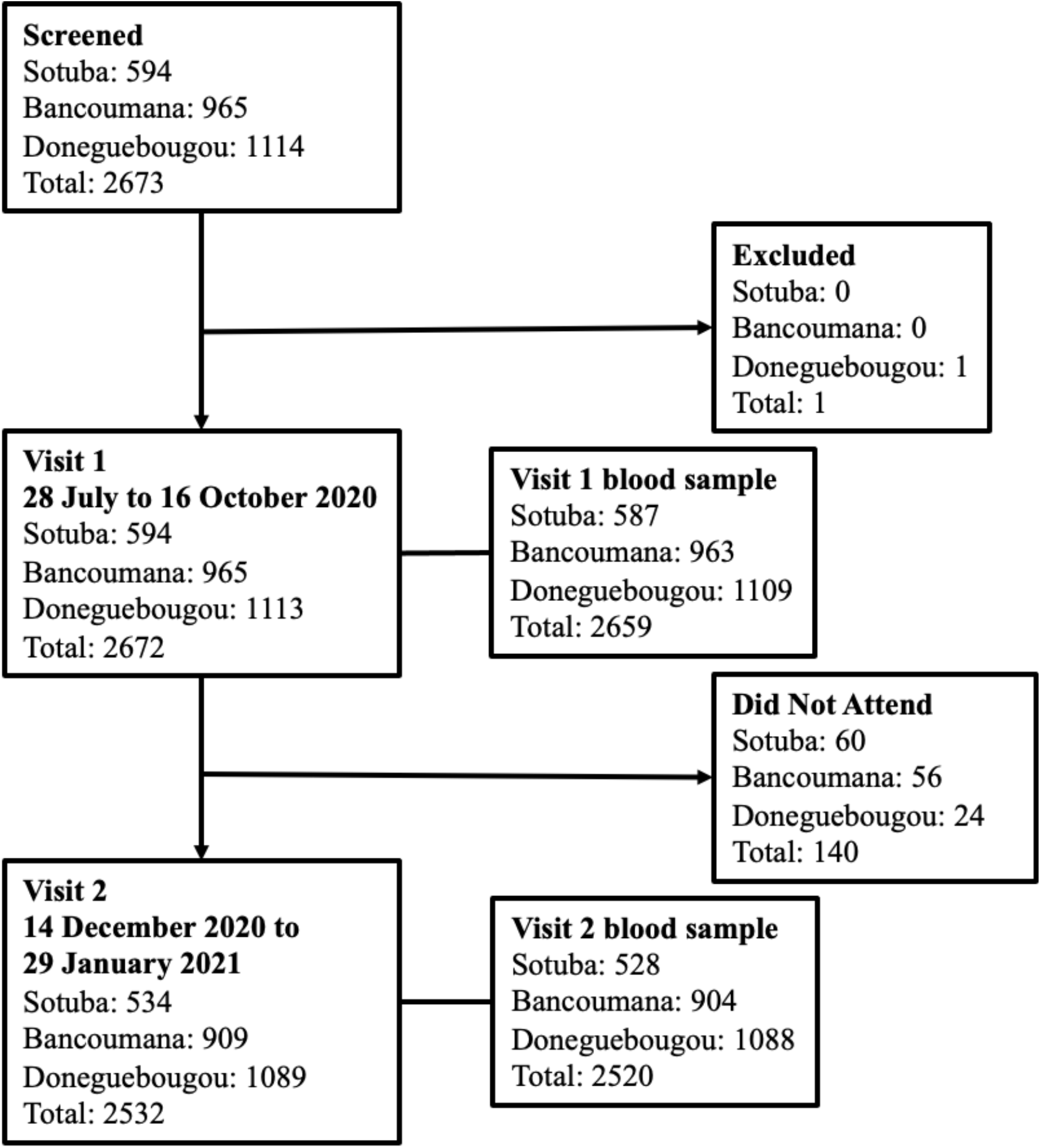
Study flow chart. Visit 1 was completed between 29 July and 16 October 2020 at the Sotuba site, 29 July and 24 September 2020 at the Bancoumana site, and 28 July and 27 August 2020 at the Donéguébougou site. Visit 2 was completed between 21 December 2020 and 26 January 2021 at the Sotuba site, 28 December 2020 and 29 January 2021 at the Bancoumana site, and 14 December 2020 and 15 January 2020 at the Donéguébougou site. A total of 94.7% (2532/2672) of participants completed visit 2.

Study population characteristics are outlined in Table 1. The study population was relatively young with very few comorbidities. At each site, a large proportion of participants were children, reflective of the age structure of the overall Malian population (Supplementary Figure 2). The median age was 14 years (IQR 8 to 31 years). No participant reported a personal history of COVID-19 diagnosis or a household member with COVID-19 diagnosis at enrollment.

**Table 1:** Study population characteristics at visit 1 (July to October 2020)

|  | **Sotuba** | **Bancoumana** | **Donéguébougou** | **Overall** |
| --- | --- | --- | --- | --- |
| **Sample size** | 594 | 965 | 1113 | 2672 |
| Individuals | 585 | 959 | 1113 | 2657 |
| Co-enrolled infants | 9 | 6 | 0 | 15 |
| **Demographics** |  |  |  |  |
| Sex, male (%) | 43.3%<br>257/594 | 51.9%<br>501/965 | 52.3%<br>582/1113 | 50.3%<br>1343/2672 |
| Age, years (median, IQR) | 14 (8-25) | 15 (8-33) | 14 (6-35) | 14 (8-31) |
| Age, years (%) |  |  |  |  |
| < 10 | 34.0%<br>202/594 | 28.9%<br>279/965 | 32.0%<br>356/1113 | 31.3%<br>837/2672 |
| 10 - 17 | 27.4%<br>163/594 | 29.4%<br>284/965 | 26.1%<br>290/1113 | 27.6%<br>737/2672 |
| ≥ 18 | 38.5%<br>229/594 | 41.7%<br>402/965 | 41.9%<br>467/1113 | 41.1%<br>1098/2672 |
| **Medical factors** |  |  |  |  |
| No co-morbid conditions (%) | 96.4%<br>564/585 | 99.2%<br>951/959 | 99.3%<br>1105/1113 | 98.6%<br>2620/2657 |
| Co-morbid conditions (%) | 3.6%<br>21/585 | 0.8%<br>8/959 | 0.7%<br>8/1113 | 1.4%<br>37/2657 |
| Obesity | 0.9%<br>5/585 | 0%<br>0/959 | 0.4%<br>5/1113 | 0.4%<br>10/2657 |
| Diabetes | 0.9%<br>5/585 | 0%<br>0/959 | 0%<br>0/1113 | 0.2%<br>5/2657 |
| HIV/other immune deficiency | 0%<br>0/585 | 0%<br>0/959 | 0%<br>0/1113 | 0%<br>0/2657 |
| Hypertension | 2.7%<br>16/585 | 1.4%<br>13/965 | 0.2%<br>2/1113 | 1.2%<br>31/2657 |
| Cardiovascular disease | 0.2%<br>1/585 | 0%<br>0/959 | 0.1%<br>1/1113 | 0.1%<br>2/2657 |
| Chronic pulmonary condition | 0%<br>0/585 | 0%<br>0/959 | 0%<br>0/1113 | 0%<br>0/2657 |
| Chronic hepatic condition | 0%<br>0/585 | 0%<br>0/959 | 0%<br>0/1113 | 0%<br>0/2657 |
| Chronic hematological condition | 0%<br>0/585 | 0.1%<br>1/959 | 0%<br>0/1113 | <0.1%<br>1/2657 |
| Chronic kidney disease | 0%<br>0/585 | 0%<br>0/959 | 0%<br>0/1113 | 0%<br>0/2657 |
| Chronic neurological impairment/disease | 0%<br>0/585 | 0%<br>0/959 | 0%<br>0/1113 | 0%<br>0/2657 |
| Malignancy | 0%<br>0/585 | 0%<br>0/959 | 0.1%<br>1/1113 | <0.1%<br>1/2657 |
| BCG vaccination (%) | 88.9%<br>520/585 | 70.2%<br>673/959 | 85.2%<br>948/1113 | 80.6%<br>2141/2657 |
| Antimalarial use (4 weeks prior to visit) (%) | 3.6%<br>21/585 | 3.9%<br>37/959 | 0.1%<br>1/1113 | 2.2%<br>59/2657 |
| Smoker (%)* | 2.9%<br>15/526 | 0.8%<br>7/904 | 4.4%<br>48/1089 | 2.8%<br>70/2519 |
| Pregnancy (%) | 1.0%<br>6/585 | 1.6%<br>15/959 | 0.2%<br>2/1113 | 0.9%<br>23/2657 |
| Trimester 1 | 0.2%<br>1/585 | 0.1%<br>1/959 | 0.2%<br>2/1113 | 0.2%<br>4/2657 |
| Trimester 2 | 0.3%<br>2/585 | 0.9%<br>9/959 | 0%<br>0/1113 | 0.4%<br>11/2657 |
| Trimester 3 | 0.5%<br>3/585 | 0.5%<br>5/959 | 0%<br>0/1113 | 0.3%<br>8/2657 |
| Post-partum (< 6 weeks) | 0.2%<br>1/585 | 0.2%<br>2/959 | 0%<br>0/1113 | 0.1%<br>3/2657 |
| **Social factors** |  |  |  |  |
| Employed in healthcare setting | 1.2%<br>7/585 | 1.8%<br>17/959 | 4.9%<br>55/1113 | 3.0%<br>79/2657 |
| Household member employed in healthcare setting | 22.6%<br>132/585 | 21.2%<br>203/959 | 1.5%<br>17/1113 | 13.2%<br>352/2657 |
| Household member previously diagnosed with COVID-19 | 0%<br>0/585 | 0%<br>0/959 | 0%<br>0/1113 | 0%<br>0/2657 |
| Household size (mean, SD) | 10.2 (5.7) | 7.1 (4.1) | 6.8 (2.9) | 7.7 (4.3) |

### Seroprevalence of SARS-CoV-2 antibodies

The adjusted seroprevalence of SARS-CoV-2 antibodies across all sites at visit 1 conducted July to October 2020 was 10.9% (95% CI: 8.1 to 13.6) and increased markedly to 54.7% (95% CI: 44.4 to 65.0) at visit 2 conducted December 2020 to January 2021 (Figure 2). For each visit, seroprevalence differed across sites (visit: 1 p<0.0001, visit 2: p<0.0001). The rate of SARS-CoV-2 antibody detection was the highest in the urban site of Sotuba and increased from 19.0% (95% CI: 14.2 to 23.8) to 70.4% (95% CI: 56.8 to 84.1) between visit 1 and visit 2 (Figure 1, Figure 2, Supplementary Table 1, Supplementary Figure 3). In the more rural Bancoumana township, antibodies against SARS-CoV-2 were detected at a lower rate, but a similar increase was observed between visit 1 and visit 2 (6.5% (95% CI: 4.1 to 9.0) to 52.1% (95% CI: 41.9 to 62.3). Seroprevalence was lowest in the rural village of Donéguébougou, but similarly increased between visit 1 and visit 2 (5.0% (95% CI: 2.8 to 7.1) to 35.0% (95% CI: 27.9 to 42.1). At each site, seroprevalence increased with age group (Supplementary Table 2). At the Sotuba site, the visit 2 seroprevalence was 77.2% (95% CI: 61.5 to 92.9) in participants aged 18 years or older. Although children aged <10 years had the lowest rate of SARS-CoV-2 antibody detection, there was evidence of increasing exposure over time in this age group at all sites.

**Figure 2:**
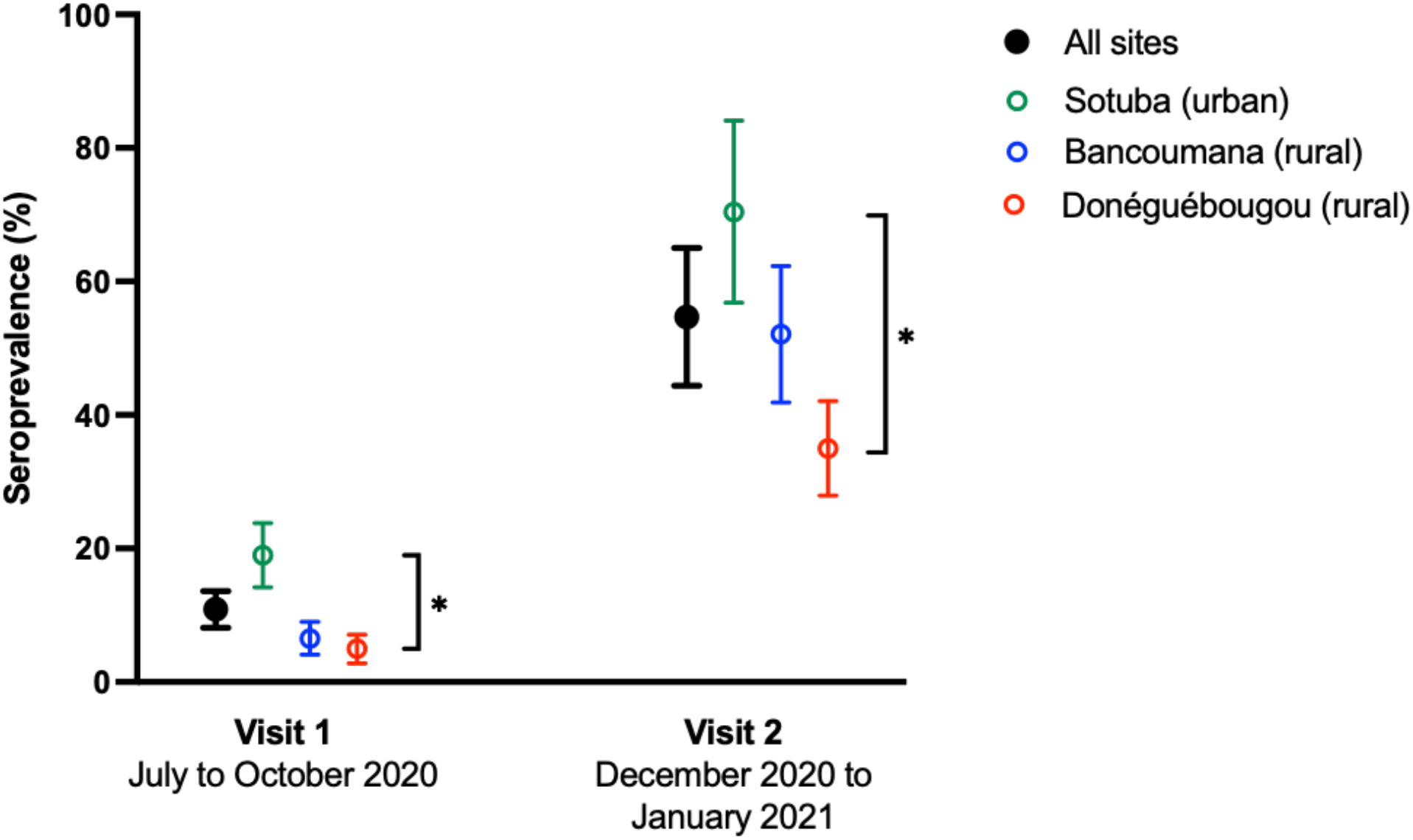
Seroprevalence of SARS-CoV-2 antibodies in Mali. Seroprevalence adjusted for population age distribution and assay sensitivity and specificity [15]. Error bars represent 95% confidence intervals. Asterisk represents p<0.0001 in comparison between sites.

**Figure 2:**
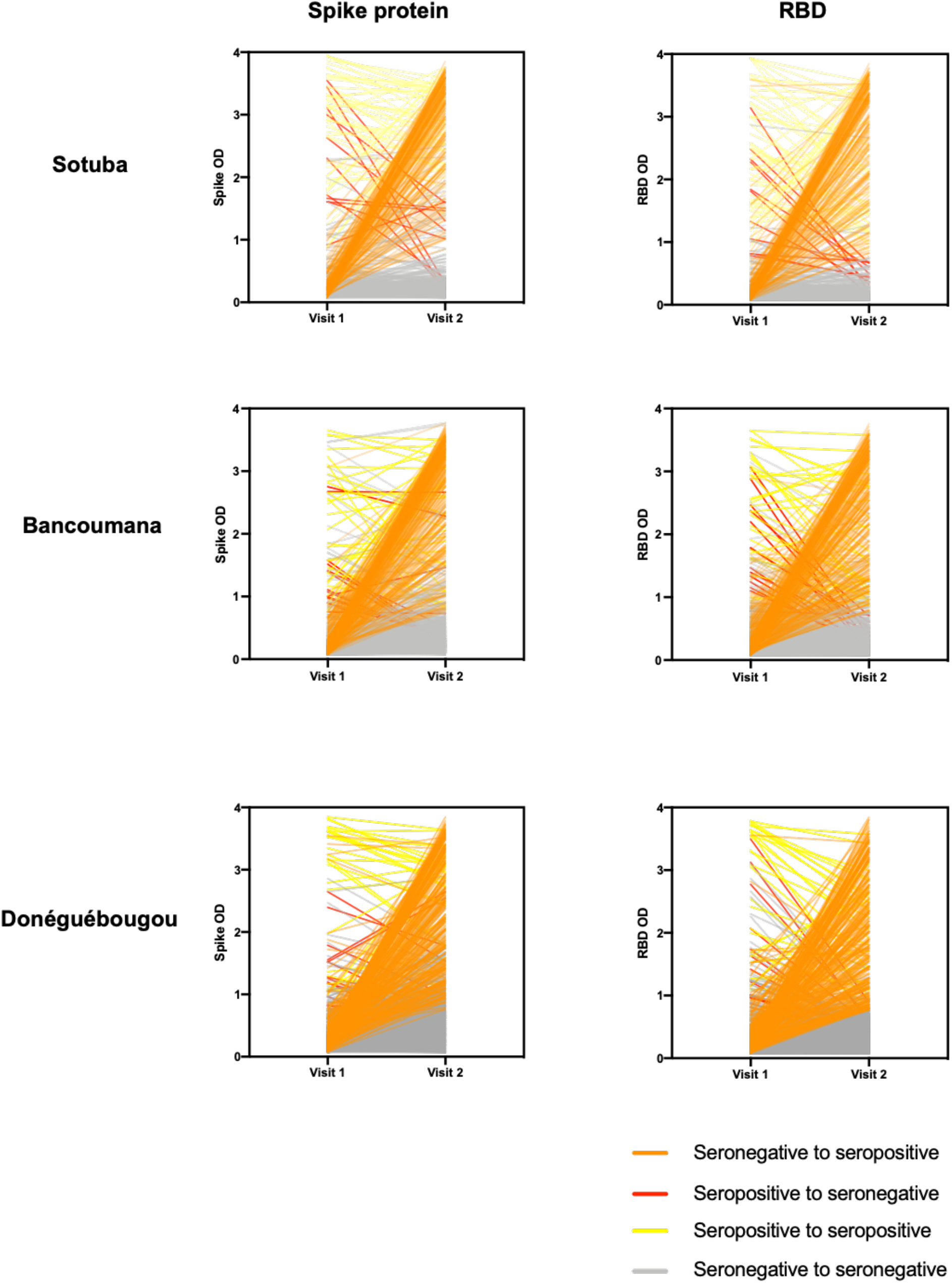
Longitudinal SARS-CoV-2 antibody reactivity spike protein and RBD at study sites: Sotuba (top row), Bancoumana (middle row) and Donéguébougou (bottom row). RBD: receptor binding domain, OD: optical density Visit 1: 28 July to 16 October 2020 Visit 2: 14 December 2020 to 29 January 2021

The cumulative adjusted SARS-CoV-2 exposure prevalence across all sites at visit 2 was 58.5% (95% CI: 47.5 to 69.4). The cumulative seroprevalence was 73.4% (95% CI: 59.2 to 87.5) at Sotuba, 53.2% (95% CI: 42.8 to 63.6) at Bancoumana, and 37.1% (95% CI: 29.6 to 44.5) at Donéguébougou (Table 2). The daily rate of infection (adjusted incidence of new cases between visit 1 and visit 2 divided by median number of days between visits) was 0.45%/day at Sotuba, 0.42%/day and Bancoumana, and 0.19%/day at Donéguébougou. The overall infection rate was 0.41%/day. This represents an infection rate of approximately 1% of the community populations every 3 days between visit 1 and visit 2.

**Table 2:** Cumulative SARS-CoV-2 exposure prevalence and rate of infection at Sotuba (urban), Bancoumana (rural town) and Donéguébougou (rural village) sites.

| Site | Visit 1 (median)<sup>1</sup> | Visit 2 (median) | Cumulative exposure prevalence Visit 2 (95% CI)<sup>2</sup> | Rate of infection (% population infected/day)<sup>3</sup> |
| --- | --- | --- | --- | --- |
| **Sotuba** | 6 August 2020<br>(29 July to 16 October 2020) | 24 December 2020<br>(21 December 2020 to 26 January 2021) | 73.4%<br>(59.2-87.5) | 0.45 |
| **Bancoumana** | 11 September 2020<br>(29 July to 24 September 2020) | 6 January 2021<br>(28 December 2020 to 29 January 2021) | 53.2%<br>(42.8-63.6) | 0.42 |
| **Donéguébougou** | 13 August 2020<br>(28 July to 27 August 2020) | 19 December 2020<br>(14 December 2020 to 15 January 2021) | 37.1%<br>(29.6-44.5) | 0.19 |

<sup>1</sup>Median visit date defined as date half of all sample collections were completed.
<sup>2</sup>Cumulative seropositivity rate calculated by adding seropositive cases from visit 1 that were seronegative at visit 2 before calculating adjusted seroprevalence.
<sup>3</sup>Adjusted incidence of new cases between visit 1 and visit 2 divided by median number of days between visits

### SARS-CoV-2 seroconversion from pre-pandemic to Visit 1 (July to August 2020)

Pre-pandemic samples were available for 402 participants from Bancoumana to evaluate seroconversion episodes (Supplementary Figure 4). In the group with pre-pandemic blood samples available, 19/402 demonstrated dual SARS-CoV-2 spike and RBD antigen seroconversion, and 19/402 were seropositive. There was a strong concordance between seroconversion and seropositivity, confirming the utility of the assay cutoffs. A total of 16/402 demonstrated both dual antigen seroconversion and seropositivity (Cohen’s kappa 0.83 (95% CI: 0.70 to 0.96)).

### Factors associated with SARS-CoV-2 serostatus

Factors associated with SARS-CoV-2 serostatus were evaluated by univariate analysis followed by multiple logistic regression for visit 1 and visit 2. Co-variates associated with serostatus at visit 1 in the univariate analysis were female sex, age, participant employment at a healthcare facility, household member employment at a healthcare facility, household size, and self-reporting of any symptoms, systemic symptoms, fever, chills, myalgia, respiratory symptoms, or gastrointestinal symptoms since March 2020 (Supplementary Table 3). Following regression, age in years (OR 1.01, 95% CI: 1.00 to 1.02), study site Sotuba (OR 2.61, 95% CI: 1.76 to 3.89), and participant employment at a healthcare facility (OR 2.47, 95% CI: 1.13 to 4.90), remained associated with seropositivity (Figure 3). Co-variates associated with serostatus at visit 2 (assessing new seropositive cases only) in the univariate analysis were age in years, household member employment at a healthcare facility, any medical comorbidity, self-reported systemic symptoms, chills, fatigue, and respiratory symptoms since visit 1 (Supplementary Table 4). Following regression, age in years (OR 1.02, 95% CI: 1.01 to 1.02), study site Sotuba (OR 1.35, 95% CI: 1.06 to 1.73), and study site Donéguébougou (OR 0.60 95% CI: 0.47 to 0.76) remained associated with serostatus (Figure 3). Among participants reporting symptoms, seeking medical attention for symptoms was associated with seropositivity (OR 1.76, 95% CI: 1.30 to 2.39) at visit 2, but not at visit 1.

**Figure 3:**
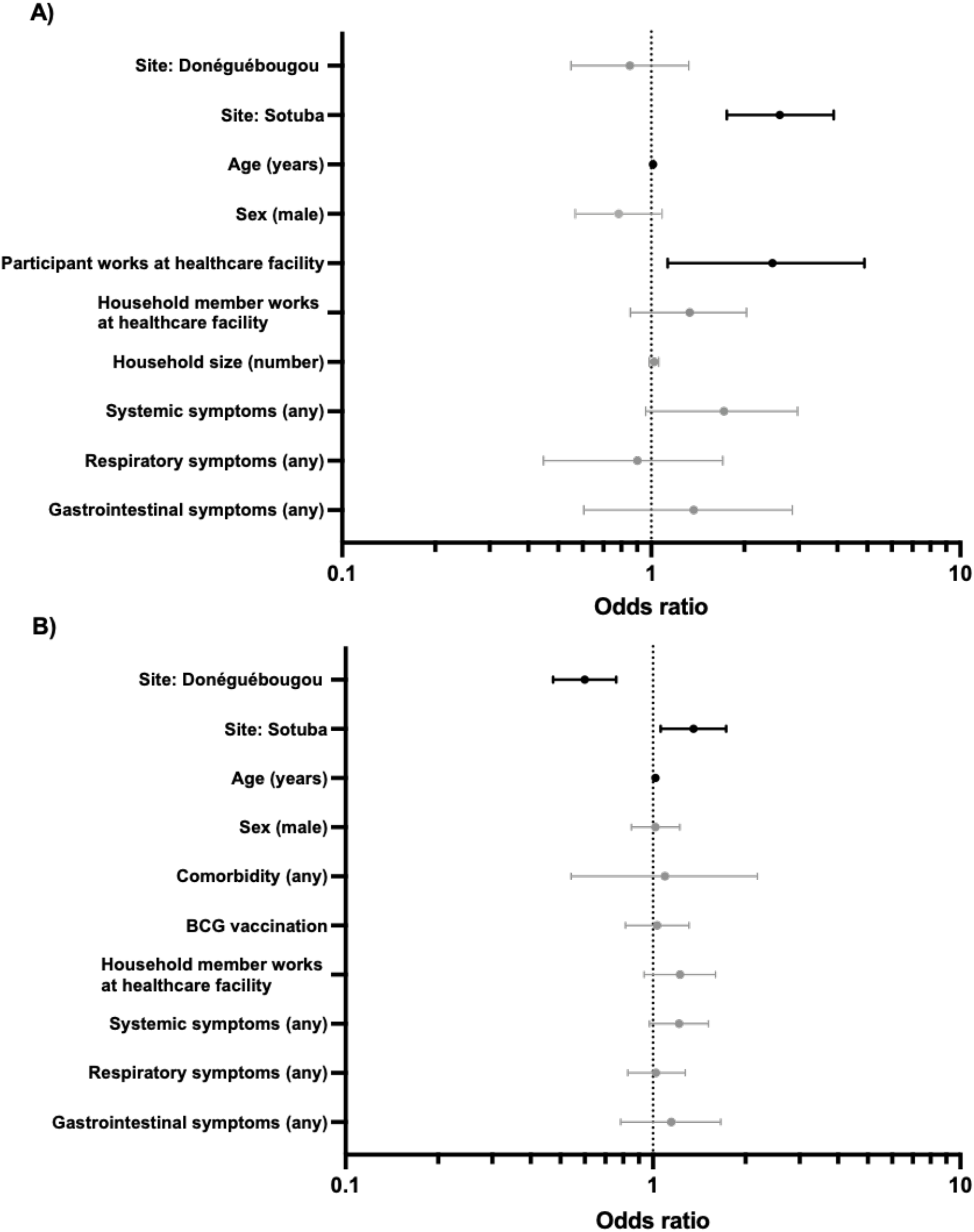
Effect of selected co-variates on seropositivity at A) visit 1 (n=2646, July to October 2020) and B) visit 2 (n=2343, December 2020 to January 2021) by multiple logistic regression. Site, age, sex and symptom categories were included *a priori*. Other co-variates were included if p<0.05 in univariate analysis (Supplementary Table 3 and 4). 173/2646 participants aged >12 months were seropositive at visit 1. 724/2343 participants aged >12 months were seropositive at visit 1. Infants aged 6-12 months underwent limited history collection and were not included in analysis.

### SARS-CoV-2 antibody dynamics between Visit 1 and Visit 2

In a longitudinal assessment of participants that were seropositive at visit 1 (July to October 2020), almost three-quarters remained seropositive at visit 2 (December 2020 to January 2021) (73.2%, 115/157) (Supplementary Table 5). The mean time between sample collections was 131.6±14.5 days. Among the 26.8% (42/157) of subjects that seroreverted, 21/42 reverted below the threshold for both spike protein and RBD, while 19/42 seroreverted RBD only, and 2/42 seroreverted spike protein only (Figure 2). In all participants seropositive at visit 1, RBD assay absorbance values waned faster than spike protein assay absorbance values at each study site (-0.47 OD units/100 days (95% CI: -0.60 to -0.34) versus -0.10 OD units/100 days (95% CI: -0.23 to +0.03). In an exploratory univariate comparison of serostable and seroreverting individuals, seroreversion was associated with male sex and smaller household size (Supplementary Table 6).

### Clinical presentation in SARS-CoV-2 seroconverters

Symptoms since the first cases of COVID-19 were detected in Mali in March 2020 were reported infrequently by seropositive participants (n=173) at visit 1 (July to October 2020). Headache (11% (19/173)) and fever (8.7%, (15/173)) were the most common. Several systemic symptoms including fever (8.7% (17/173) vs 4.0% (100/2473), p=0.0101), chills (1.7% (3/173) vs 0.4% (9/2473), p=0.0390), myalgia (3.5% (6/173) vs 0.7% (18/2473), p=0.0036), and headache (11.0% (19/173) vs 3.7% (91/2473), p<0.0001) were more frequently reported by seropositive participants compared to seronegative participants (Supplementary Table 3). Grouped systemic symptoms (any) were almost independently associated with serostatus following multiple logistic regression (OR 1.72, 95% CI: 0.96-2.98, Figure 3).

There was a larger group of newly seropositive participants at visit 2 to assess (n=724). Symptoms occurring between visit 1 (July to October 2020) and visit 2 (December 2020 to January 2021) were reported with a greater frequency compared to the prior reporting period (Supplementary Table 3 and 4). The most common symptoms were rhinorrhea (26.1% (189/724)), headache (22.7% (164/724)), cough (19.1% (138/724)) and fever (9.9% (72/724)). The remaining symptoms were reported in less than 5% of individuals, including loss of smell or taste (2.2% (16/724)). Among seropositive participants, 48.6% (352/724) reported a history of any symptoms, compared to 49.3% (803/1629) in seronegative participants. As a result of increased background illness rates, it is difficult to establish which symptoms are associated with COVID-19 in the study population. Chills (3.7% (27/724) vs 2.1% (34/1619), p=0.0250) and fatigue (4.3% (31/724) vs 2.5% (41/1619), p=0.0275) were more frequently reported by seropositive participants compared to seronegative participants. Similar to the assessment at visit 1, grouped systemic symptoms (any) were almost independently associated with serostatus following multiple logistic regression (OR 1.22, 95% CI: 0.97-1.52, Figure 3) In seropositive participants reporting symptoms, 15.6% (55/352) reported absenteeism from work or school, 63.4% (223/352) reported seeking medical attention, and 0.9% (3/352) reported hospitalization. The three participants reporting hospitalization were a two year old male with fever, cough and rhinorrhea, a 12 year old male with a headache, and a 30 year old male with fever, headache and rhinorrhea. In participants reporting symptoms at visit 2, seropositive individuals were more likely to have sought medical attention for symptoms compared to seronegative individuals (63.4% (223/352) vs 45.9% (366/797), p<0.0001, Supplementary Table 4).

In a subset of 146 healthy adult participants co-enrolled in a Phase 2 clinical trial at the Bancoumana site [17] during this SARS-CoV-2 seroprevalence study, clinical trial adverse events occurring between visit 1 and 7 days before visit 2 were assessed to better understand the clinical presentation of COVID-19. There was a low frequency of most clinical and laboratory adverse events irrespective of serostatus (Supplementary Table 7). Pain (non-specific) was more common in the newly seropositive group compared to the seronegative group (8.2% (5/61) vs 0% (0/85), p=0.0139). There was no statistically significant difference in the frequency of potentially COVID-19 related adverse events including bronchitis (1.6% (1/61) vs 3.5% (3/85), p=0.6403), cough (3.3% (2/61) vs 1.2% (1/85), p=0.5714), pyrexia (1.6% (1/61) vs 0% (0/85), p=0.4296), chills (1.6% (1/61) vs 0% (0/85), p=0.4218), headache (11.5% (7/61) vs 15.3% (13/85), p=0.6280), rhinitis (34.4% (21/61) vs 23.5% (20/85), p=0.2619), sinobronchitis (3.3% (2/61) vs 3.5% (3/85), p>0.9999), leukopenia (9.8% (6/61) vs 4.7% (4/85), p=0.2020), or thrombocytopenia (3.3% (2/61) vs 0% (0/85), p=0.1729). There was no difference in the grading of the most commonly reported adverse events between the seropositive group and the seronegative group (Supplementary Table 8).

Similarly, in a subset of 1037 participants of all ages co-enrolled in a community Phase 2 clinical trial at the Donéguébougou [18], there was a low frequency of most clinical and laboratory adverse events irrespective of serostatus (Supplementary Table 9). Headache (18.3% (46/252) vs 9.4% (74/785), p=0.0003) and rhinitis (33.3% (84/252) vs 25.1% (197/785), p=0.0116) were more common in the newly seropositive group compared to the seronegative group. There was no statistically significant difference in the frequency of other potentially COVID-19 related adverse events. Dental caries (3.2% (8/252) vs 1.1% (9/785), p=0.0416) and gastritis (1.2% (3/252) vs 0.1% (1/785), p=0.0466) were also observed more frequently in the seroconverting group. There was no difference in the grading of the most commonly reported adverse events between the seropositive group and the seronegative group, including headache and rhinitis (Supplementary Table 10).

## DISCUSSION

In this prospective cohort study of three sites in urban and rural Mali, we provide evidence of marked community transmission between July 2020 and January 2021 using longitudinal population serosurveillance. Seroprevalence estimates using a two-antigen ELISA with cutoffs adapted to improve specificity in the study population identified an infection rate of approximately 1% of the population every three days in the study communities between visits, and a cumulative SARS-CoV-2 exposure rate of 58.5% (95% CI: 47.5 to 69.4). Cumulative exposure varied between sites, and was 73.4% (95% CI: 59.2 to 87.5) in the urban community of Sotuba, 53.2% (95% CI: 42.8 to 63.6) in the rural town of Bancoumana, and 37.1% (95% CI: 29.6 to 44.5) in the rural village of Donéguébougou in January 2021. No study participant reported a history of COVID-19 diagnosis in our study. Previously, serosurveillance in the US has estimated a case detection ratio of 20%, while in Zambia, combined rt-PCR/serosurveillance estimated a 1% case detection ratio [10, 19]. In our study, these seroprevalence findings would suggest a case detection ratio of approximately 0.1-0.2% in Mali based on the number of previously reported cases nationwide [20]. This highlights the need for improved access to diagnostic testing in the community to better understand the pandemic, although targeting these diagnostics will remain a challenge in the presence of limited disease. The dearth of apparent COVID-19 disease in our study population is consistent with the pandemic epidemiology previously reported in sub-Saharan Africa [1, 2]. Our study included a large number of children and is reflective of the population age structure in the region.

Based on the estimated rate of infection in our study, the hypothetical ‘herd immunity’ threshold of 70-80% may have been reached among adults in Sotuba (77.2% (95% CI: 61.5 to 92.9). It is unclear if the evidence of natural infection rates provided by serosurveillance can be used to approximate level of population protection although the presence of SARS-COV-2 antibodies has been associated with an approximately 80% lower risk of infection compared to seronegative individuals in adult populations [21, 22]. The clinical significance of our assay is uncertain, however we have previously shown strong correlation between spike protein and RBD ELISA assay absorbance and pseudovirus neutralization activity in US samples [6]. In our study, there was relatively faster waning of RBD absorbance values over time compared to spike protein. Despite the relatively mild illness reported, the durability of spike protein IgG antibodies suggests a relatively long-lasted humoral response in our study population, and seropositivity may be a surrogate marker for longer term cellular immunity [23]. Taken together, the rapid increase in SARS-CoV-2 antibody seroprevalence and limited attributable severe illness during the study period may reflect a degree of protection in the community.

Conversely, the widespread SARS-CoV-2 transmission suggested by the marked increase in seroprevalence during our study could promote the emergence of new variants that may affect any natural herd immunity. In a similar high seroprevalence scenario in Manaus, Brazil, a large resurgence in cases was reported following introduction of the B.1.1.248 variant, suggesting limited cross-protection from prior infection [24, 25]. Notably, that study employed a lowered single antigen cutoff for seropositivity, which may have overestimated population exposure [25]. We are uncertain of the locally circulating variants in Mali, and whether our study may have coincided with emergence of a new variant. The South African variant B.1.351 was reported in Ghana in early January 2021 [26], and became the dominant circulating strain in South Africa over the study period [27].

In our study population, seropositivity early in the pandemic was associated with increasing age, employment in a healthcare setting, and residence in the urban community of Sotuba. Seropositivity later in the pandemic was associated only with increasing age and residence in the urban community of Sotuba. Residence in the rural village of Donéguébougou was associated with relative protection from infection. This is consistent with other reports, where there is marked regional variation in seroprevalence rates, children demonstrate lower seropositivity rates compared to adults, and healthcare workers demonstrate higher seropositivity rates compared to the wider community [8, 10]. Symptom history was not reliably associated with serostatus, highlighting the relatively limited clinical burden of the pandemic in the study population and the challenge of detecting cases by passive surveillance.

The proportion of participants reporting a history of symptoms in our population is in keeping with smaller SARS-CoV-2 serosurveys conducted in West Africa [8, 9]. Between July 2020 and January 2021 there was a high rate of background illness, where 49.1% (1155/2353) of participants reported symptoms irrespective of serostatus. This time period coincides with local seasonal malaria and emphasizes the importance of readily available and reliable diagnostics, particularly in regions where the differential diagnosis for non-specific symptoms is broad and may include malaria and viral hemorrhagic fevers. The lack of excess clinical illness in the SARS-CoV-2 seroconverting group, including in a subpopulation intensively followed up for adverse events, suggests that the rate of COVID-19 attributable symptoms is low in our population, especially during the malaria transmission season. In an active surveillance study in Zambia, 23.8% of PCR-confirmed COVID-19 cases reported symptoms [10], which falls within the background rate of symptoms in our study.

While our study is not powered to determine if severe COVID-19 is less common in sub-Saharan compared to other settings, we found minimal difference in reported symptoms, hospitalization or absenteeism based on serostatus. Using estimated COVID-19 hospitalization rates from the United States adjusted for the age-structure of our study population, we would expect a 2% hospitalization rate, or approximately 30 hospitalization events among the nearly 1500 SARS-CoV-2 infections estimated in our study [28]. In total, six hospitalizations were reported among the study population, three in seropositive participants and three in seronegative participants. Seeking medical attention for reported symptoms was more frequent among participants seroconverting between visit 1 and visit 2, suggesting that there may be an opportunity to identify some cases acutely if diagnostic resources are available. As our study population was able to attend the study site clinic to access care during the course of the study, it is uncertain if this rate of seeking medical attention is reflective of the wider population, however in the Zambian study 65.1% of symptomatic individuals reported seeking medical attention [10].

These data provide valuable information for use in the Malian Public Health effort. The longitudinal clinical and laboratory data collected in our study helps to close the gap of community-based pandemic data in the West African region, and may assist in ongoing Public Health interventions, including the design of vaccination programs. In this study, two visits were completed by 94.8% (2533/2672) of participants, including a large number of children, which is reflective of the local population age structure. The two-antigen ELISA used in this study has been used for population seroprevalence in the United States [19], and has been optimized to improve specificity in Mali [6]. In this study, there was a strong concordance between two-antigen seropositivity, and two-antigen four-fold seroconversion compared to baseline in a subset of 402 participants (Cohen’s kappa 0.834 (95% CI: 0.704 to 0.964)), providing further reassurance of assay performance.

Our study has several limitations, including a sample size that is not sufficient to determine the rate of uncommon severe outcomes in the community, and the risk of recall bias in reported symptoms history. In our study, reported symptoms history was similar in the overall study population to adverse events recorded for a subpopulation co-enrolled in parallel clinical trials. Furthermore we would expect recall bias to be similar between seropositive and seronegative participants. The study population was not selected randomly, and therefore there is also a risk of selection bias. Notably, no participant reported a personal history of prior COVID-19 diagnosis or household member with a diagnosis at enrollment, and there was a high proportion of community participation, particularly at the Donéguébougou site.

The evidence here of high community seroprevalence can inform prioritization and implementation of vaccination programs in populations with a young age structure and limited evidence of substantial clinical disease. Information on circulating virus variants will also be valuable to decision-making, as efficacy of any available vaccines may vary against locally circulating variants.

This study provides further evidence that Africa has not been spared by SARS-CoV-2, and that the epidemiology of disease in Malian communities may be primarily subclinical and within background illness rates. In this setting, community mitigation strategies may differ to other regions, and ongoing surveillance and augmentation of diagnostics, including characterizing locally circulating variants will be critical to implement and monitor an effective vaccination program.

## Data Availability

Data referred to in the manuscript are included in the text and associated supplementary materials.

## ACKNOWLEDGEMENTS

We gratefully acknowledge the support of Dr. Thayne Dickey (LMIV/NIAID) for providing positive control monoclonal neutralizing antibody CR3022; Rathy Mohan (LMIV/NIAID) for database support; Dr David Cook and Alemush Imeru for curation of the MEDDra coded adverse event data; Dr Rebecca Prevots (LCIM/NIAID) and Dr Dean Follman (BRB/NIAID) for assistance designing collection and analysis of study data; Patrick Gorres (LMIV/NIAID) for editorial assistance; the Malian COVID-19 Coordinator; and Ministry of Health for permission to partner in developing serosurveillance capacity in Mali.

## DATA AVAILABILTY STATEMENT

De-identified data collected for this study may be made available to others after approval of approval and with a signed data access agreement.

