## Supplementary Tables and Figures for "Rapidly increasing SARS-CoV-2 seroprevalence and limited clinical disease in three Malian communities: a prospective cohort study"

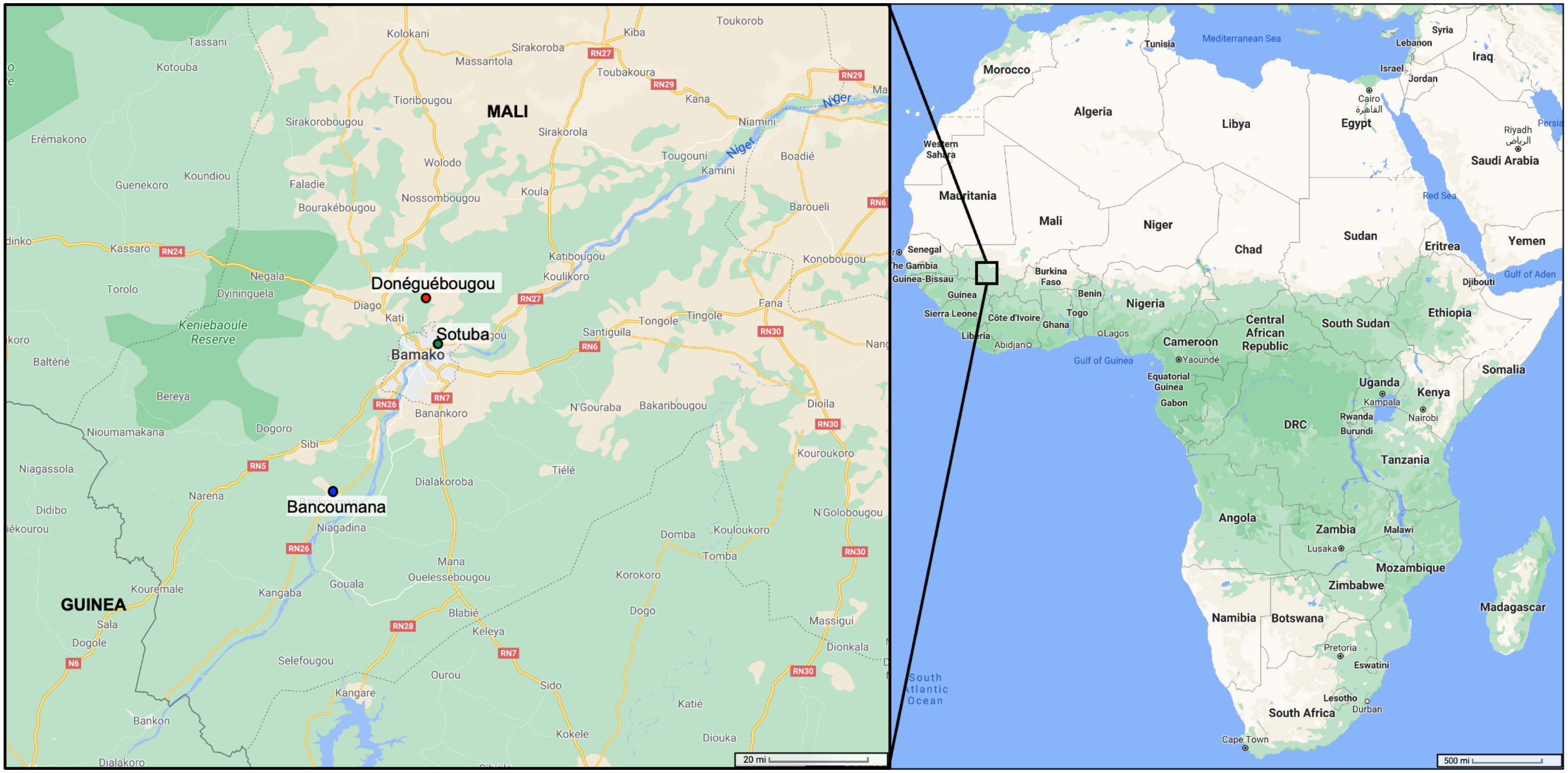

**Supplementary Figure 1: Map of study sites**

Adapted from Google Maps

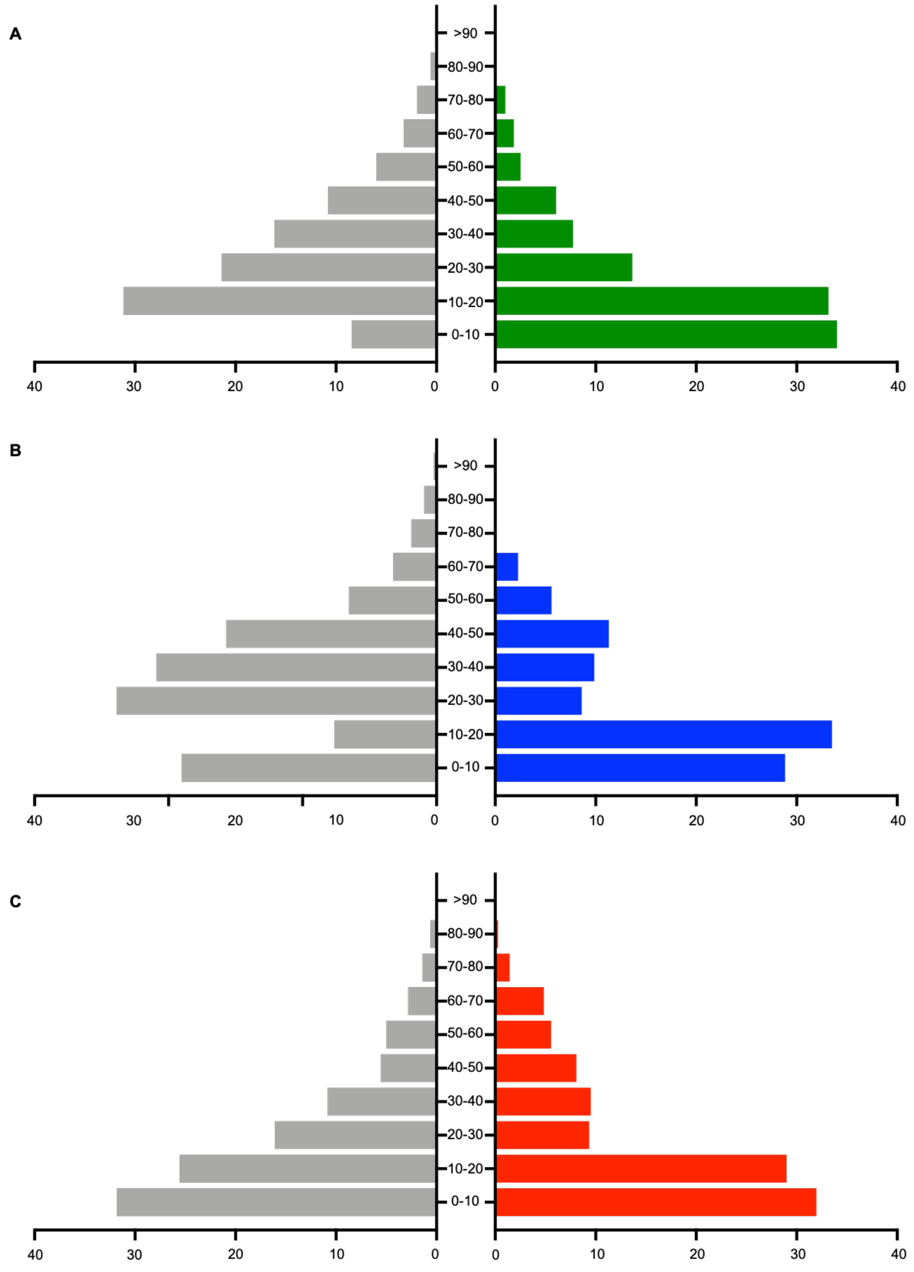

**Supplementary Figure 2: Age structure of A) Sotuba (urban), B) Bancoumana (rural), and C) Donéguébougou (rural). Age group in years (y-axis) versus percentage of overall population (x-axis).**
Grey histograms represent census data for each study site.
Colored histograms represent sample for each study site.

**Supplementary Table 1: Seroprevalence of SARS-CoV-2 antibodies at visit 1 and visit 2 at Sotuba (urban), Bancoumana (rural town) and Donéguébougou (rural) sites**.

| **Site** | **Dates of sample collection** | **Crude seropositivity rate (95% CI)** | **Adjusted seropositivity rate (95% CI)[1]** |
|---|---|---|---|
| Sotuba (visit 1) N=587 | 29 July to 16 October 2020 | 13.1% (10.4-15.9) | 19.0% (14.2-23.8) |
| Sotuba (visit 2) N=528 | 21 December 2020 to 26 January 2021 | 44.9% (40.7-49.1) | 70.4% (56.8-84.1) |
| Bancoumana (visit 1) N=963 | 29 July to 24 September 2020 | 5.3% (3.9-6.7) | 6.5% (4.1-9.0) |
| Bancoumana (visit 2) N=904 | 28 December 2020 to 29 January 2021 | 35.5% (32.4-38.6) | 52.1% (41.9-62.3) |
| Donéguébougou (visit 1) N=1109 | 28 July to 27 August 2020 | 4.1% (2.9-5.2) | 5.0% (2.8-7.1) |
| Donéguébougou (visit 2) N=1088 | 14 December 2020 to 15 January 2021 | 25.8% (23.2-28.4) | 35.0% (27.9-42.1) |

[1]Adjusted for population age distribution and assay sensitivity and specificity [1].

1. Lang, Z. and J. Reiczigel, *Confidence limits for prevalence of disease adjusted for estimated sensitivity and specificity.* Prev Vet Med, 2014. **113**(1): p. 13-22.

**Supplementary Table 2: Age-stratified seroprevalence of SARS-CoV-2 antibodies at visit 1 and visit 2 at Sotuba (urban), Bancoumana (rural town) and Donéguébougou (rural) sites.**

| Site | Dates of sample collection | Crude seropositivity rate (95% CI) | | | Adjusted seropositivity rate (95% CI)[1] | | |
|---|---|---|---|---|---|---|---|
| | | <10 years | 10-17 years | ≥18 years | <10 years | 10-17 years | ≥18 years |
| Sotuba (visit 1) N=587 | 29 July to 16 October 2020 | 10.7% (6.3-15.1) | 11.7% (6.6-16.7) | 16.2% (11.4-21.0) | 13.8% (8.4-19.2) | 15.1% (9.0-21.1) | 21.3% (14.8-27.8) |
| Sotuba (visit 2) N=528 | 21 December 2020 to 26 January 2021 | 29.1% (22.4-35.7) | 47.3% (39.4-55.2) | 57.2% (50.4-64.0) | 38.8% (28.9-48.7) | 63.7% (49.5-77.9) | 77.2% (61.5-92.9) |
| Bancoumana (visit 1) N=963 | 29 July to 24 September 2020 | 4.0% (1.5-6.4) | 6.0% (3.1-8.8) | 5.7% (3.4-8.1) | 4.6% (1.6-7.6) | 7.3% (3.8-10.9) | 7.0% (3.9-10.1) |
| Bancoumana (visit 2) N=904 | 28 December 2020 to 29 January 2021 | 24.2% (19.1-29.4) | 36.6% (30.8-42.4) | 42.6% (37.6-47.6) | 32.3% (24.2-40.3) | 49.2% (38.3-60.0) | 57.3% (45.6-69.0) |
| Donéguébougou (visit 1) N=1109 | 28 July to 27 August 2020 | 3.1% (1.2-5.1) | 1.7% (0.0-3.5%) | 6.2% (4.0-8.5) | 3.4% (0.9-6.0%) | 1.5% (0-3.9) | 7.7% (4.6-10.8) |
| Donéguébougou (visit 2) N=1088 | 14 December 2020 to 15 January 2021 | 12.2% (8.8-15.7) | 28.5% (23.2-33.7) | 34.7% (30.4-39.1) | 15.8% (11.0-20.7) | 41.0% (31.5-50.5) | 46.6% (36.8-56.3) |

[1]Adjusted for assay sensitivity and specificity [1].

1. Lang, Z. and J. Reiczigel, *Confidence limits for prevalence of disease adjusted for estimated sensitivity and specificity.* Prev Vet Med, 2014. **113**(1): p. 13-22.

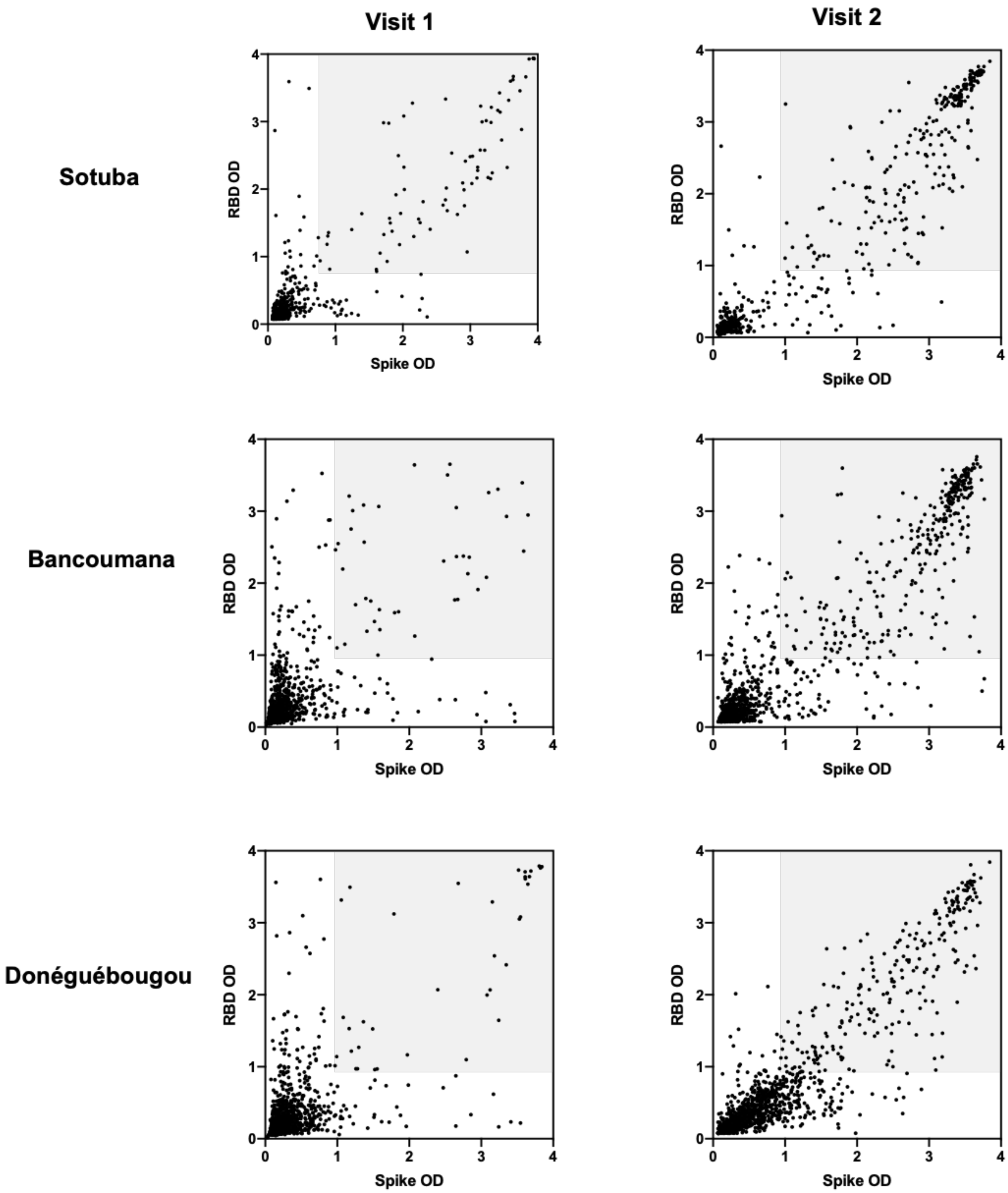

**Supplementary Figure 3: SARS-CoV-2 antibody reactivity to spike protein and RBD over time at study sites: Sotuba (top row), Bancoumana (middle row) and Donéguébougou (bottom row)**.
RBD: receptor binding domain, OD: optical density
Visit 1: 28 July to 16 October 2020
Visit 2: 14 December 2020 to 29 January 2021
Shaded region represents ELISA measurements that exceed cutoffs to define seropositive cases

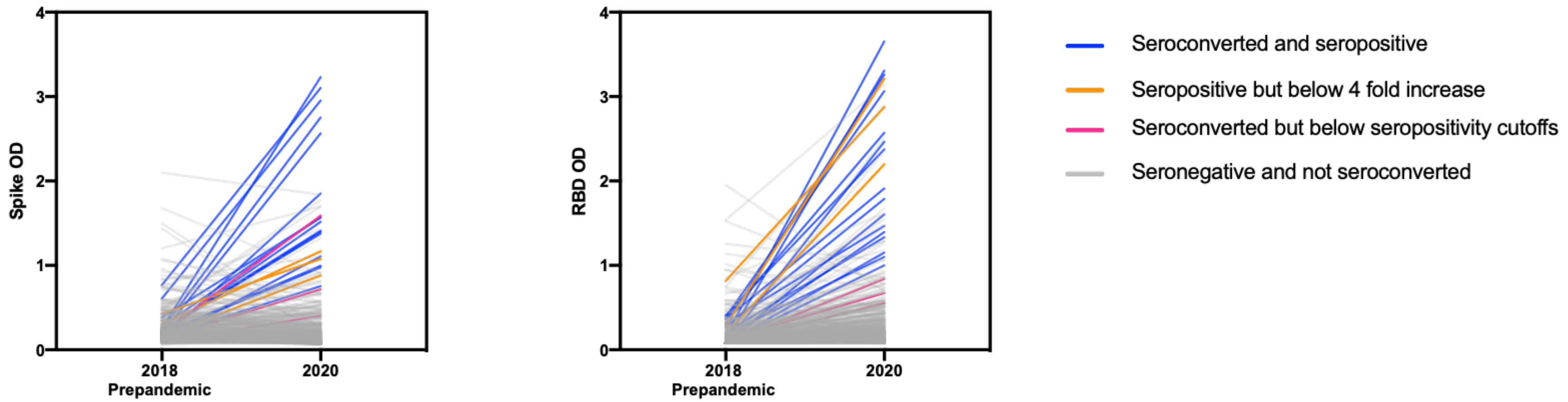

**Supplementary Figure 4: Seroconversion and seropositivity of SARS-CoV-2 antibodies to spike protein and RBD in 402 participants with pre-pandemic blood samples from Bancoumana**

RBD: receptor binding domain, OD: optical density

Seroconversion: fourfold increase in spike protein and RBD OD value from pre-pandemic sample

Seropositive: spike protein and RBD OD value above cutoff

**Supplementary Table 3: Univariate comparison of seronegative and seropositive subpopulations at visit 1 (n=2659, July/October 2020)**

| | Seronegative | Seropositive | p-value |
|---|---|---|---|
| Sample size | 2486 | 173 | |
| Co-enrolled infants | 13 | 0 | |
| **Demographics** | | | |
| Sex, male (%, n/N) | 50.9% (1266/2486) | 41.6% (72/173) | **0.0184** |
| Age, years (median, IQR) | 14 (8-31) | 18 (10-36) | **0.0066** |
| Age group (%, n/N) | | | |
| <10 years | 31.5% (783/2486) | 24.9% (43/173) | |
| 10-17 years | 28.0% (696/2486) | 23.7% (41/173) | |
| >/=18 years | 40.5% (1007/2486) | 51.4% (89/173) | |
| **Medical factors (% (n/N)** | | | |
| Any comorbidity | 1.5% (37/2473) | 2.9% (5/173) | 0.1929 |
| Pregnancy (any stage) | 0.8% (21/2473) | 1.2% (2/173) | 0.6595 |
| Smoking | 2.8% (66/2343) | 2.5% (4/157) | >0.9999 |
| Antimalarial use | 2.1% (52/2473) | 3.5% (6/173) | 0.2726 |
| BCG administration | 80.5% (1992/2473 | 79.8% (138/173) | 0.7668 |
| **Social factors** | | | |
| Works at healthcare facility | 2.8% (69/2473) | 5.8% (10/173) | **0.0352** |
| Household member works at healthcare facility | 12.8% (317/2473) | 19.7% (34/173) | **0.0145** |
| Household size (mean, SD) | 7.6 (4.2) | 8.6 (5.4) | **0.0043** |
| **Symptoms (%, n/N)** | | | |
| No symptoms since onset pandemic | 91.2% (2242/2486) | 79.2% (137/173) | |
| Symptoms since March 2020 (any) | 9.8% (244/2486) | 20.8% (36/173) | **<0.0001** |
| **Systemic symptoms (any)** | 6.4% (159/2473) | 14.5% (25/173) | **0.0003** |
| Fever | 4.0% (100/2473) | 8.7% (15/173) | **0.0101** |
| Chills | 0.4% (9/2473) | 1.7% (3/173) | **0.0390** |
| Fatigue | 0.7% (17/2473) | 1.2% (2/173) | 0.3557 |
| Myalgia | 0.7% (18/2473) | 3.5% (6/173) | **0.0036** |
| Headache | 3.7% (91/2473) | 11.0% (19/173) | **<0.0001** |
| **Respiratory symptoms (any)** | 4.5% (112/2473) | 8.1% (14/173) | **0.0413** |
| Sore throat | 0.4% (9/2473) | 1.2% (2/173) | 0.1587 |
| Cough | 2.4% (59/2473) | 4.1% (7/173) | 0.1989 |
| Rhinorrhea | 3.6% (89/2473) | 5.2% (9/173) | 0.2932 |
| Dyspnea | 0% (0/2473) | 0% (0/173) | >0.9999 |
| Wheezing | 0% (0/2473) | 0% (0/173) | >0.9999 |
| Loss of smell/taste | 0.2% (4/2473) | 0.6% (1/173) | 0.2871 |
| Other respiratory symptoms | <0.1% (1/2473) | 0% (0/173) | >0.9999 |
| **Gastrointestinal symptoms (any)** | 2.7% (67/2473) | 5.8% (10/173) | **0.0313** |
| Nausea/vomiting | 1.5% (37/2473) | 2.9% (5/173) | 0.1929 |
| Abdominal pain | 1.3% (31/2473) | 2.9% (5/173) | 0.0819 |
| Diarrhea | 0.6% (16/2473) | 0.6% (1/173) | >0.9999 |
| **Symptom severity (%, n/N)[1]** | | | |
| Missed work or school | 39.2% (93/237) | 36.1% (13/36) | 0.8547 |
| Sought medical attention | 49.4% (117/237) | 33.3% (12/36) | 0.0765 |
| Hospitalized[2] | 0.4% (1/237) | 0% (0/36) | >0.9999 |
| Duration of symptoms (any) (mean, SD) | 6.4 (8.2) | 3.6 (3.0) | 0.1126 |
| Symptomatic at visit 1 | 21.1% (50/237) | 36.1% (13/36) | 0.0564 |

[1]Indices of symptom severity collected in participants reporting any symptoms. Details were not collected from infants aged 6-12 months.
[2]Reported hospitalization: seronegative case: 8 year old female with fever, and nausea and vomiting.

**Supplementary Table 4: Univariate comparison of seronegative and new seropositive subpopulations at visit 2 (n=2353, July/October 2020 to December 2020/January 2021)**

| | Seronegative[1] | Seropositive[2] | p-value |
|---|---|---|---|
| Sample size | 1629 | 724 | |
| Co-enrolled infants | 10 | 0 | |
| **Demographics** | | | |
| Sex, male (%, n/N) | 51.7% (843/1629) | 49.6% (359/724) | 0.3482 |
| Age, years (median, IQR) | 12 (7-28) | 18 (11-27) | **<0.0001** |
| Age group (%, n/N) | | | |
| <10 years | 37.7% (614/1629) | 18.5% (134/724) | |
| 10-17 years | 26.5% (432/1629) | 30.7% (222/724) | |
| >/=18 years | 35.8% (583/1629) | 50.8% (368/724) | |
| Days between enrollment and follow up (mean, SD) | 128.1 (13.1) | 129.1 (19.8) | 0.1686 |
| **Medical factors (% (n/N)** | | | |
| Any comorbidity | 1.1% (17/1619) | 2.2% (16/724) | **0.0358** |
| Pregnancy (any stage) | 0.6% (10/1629) | 1.2% (9/724) | 0.1361 |
| Smoking | 3.2% (51/1619) | 2.1% (15/724) | 0.1763 |
| Antimalarial use | 2.0% (32/1619) | 1.9% (14/724) | >0.9999 |
| BCG administration | 81.5% (1320/1619) | 77.2% (559/724) | **0.0159** |
| **Social factors** | | | |
| Works at healthcare facility | 3.2% (52/1619) | 2.2% (16/724) | 0.2302 |
| Household member works at healthcare facility | 11.2% (181/1619) | 16.0% (116/724) | **0.0015** |
| Household size (mean, SD) | 7.6 (4.2) | 7.5 (4.3) | 0.8194 |
| **Symptoms (%, n/N)** | | | |
| No symptoms since visit 1 | 50.7% (826/1629) | 51.4% (372/724) | |
| Symptoms since visit 1 (any) | 49.3% (803/1629) | 48.6% (352/724) | 0.7887 |
| **Systemic symptoms (any)** | 22.7% (368/1619) | 27.8% (201/724) | **0.0092** |
| Fever | 8.3% (134/1619) | 9.9% (72/724) | 0.2064 |
| Chills | 2.1% (34/1619) | 3.7% (27/724) | **0.0250** |
| Fatigue | 2.5% (41/1619) | 4.3% (31/724) | **0.0275** |
| Myalgia | 2.4% (39/1619) | 2.9% (21/724) | 0.4817 |
| Headache | 19.1% (309/1619) | 22.7% (164/724) | 0.0512 |
| **Respiratory symptoms (any)** | 36.9% (598/1619) | 32.3% (234/724) | **0.0317** |
| Sore throat | 3.0% (48/1619) | 2.8% (20/724) | 0.8942 |
| Cough | 21.6% (349/1619) | 19.1% (138/724) | 0.1860 |
| Rhinorrhea | 29.7% (481/1619) | 26.1% (189/724) | 0.0752 |
| Dyspnea | 0.2% (4/1619) | 0.3% (2/724) | >0.9999 |
| Wheezing | 0% (0/1619) | 0% (0/724) | >0.9999 |
| Loss of smell/taste | 1.3% (21/1619) | 2.2% (16/724) | 0.1083 |
| Other respiratory symptoms | 0.4% (6/1619) | 0.1% (1/724) | 0.4480 |
| **Gastrointestinal symptoms (any)** | 5.6% (90/1619) | 7.0% (51/724) | 0.1878 |
| Nausea/vomiting | 2.3% (38/1619) | 3.7% (27/724) | 0.0758 |
| Abdominal pain | 3.5% (56/1619) | 5.0% (36/724) | 0.0850 |
| Diarrhea | 1.1% (17/1619) | 0.7% (5/724) | 0.4927 |
| **Symptom severity (%, n/N)[3]** | | | |
| Missed work or school | 12.5% (100/797) | 15.6% (55/352) | 0.1611 |
| Sought medical attention | 45.9% (366/797) | 63.4% (223/352) | **<0.0001** |
| Hospitalized[4] | 0.3% (2/797) | 0.9% (3/352) | 0.1711 |
| Duration of symptoms (any) (mean, SD) | 4.5 (6.2) | 4.4 (3.1) | 0.190 |
| Symptomatic at visit 2 | 38.3% (305/797) | 32.1% (113/352) | **0.0463** |

[1]Seronegative refers to seronegative individuals at visit 1 and visit 2

[2]Seropositive refers to new seropositive individuals at visit 2 (seronegative at visit 1)
[3]Indices of symptom severity collected in participants reporting any symptoms. Details were not collected from infants aged 6-12 months.
[4]Reported hospitalizations: seronegative cases: 22 year old female with fever, headache, nausea and vomiting, and abdominal pain, 14 year old male with cough. Seropositive cases: 2 year old male with fever, cough and rhinorrhea, 12 year old male with headache, and 30 year old male with fever, headache and rhinorrhea.

**Supplementary Table 5: Longitudinal assessment of participants seropositive at visit 1 (n=157, July/October 2020 to December 2021/January 2021)**

| | **Sotuba** | **Bancoumana** | **Donéguébougou** | **Overall** |
|---|---|---|---|---|
| Sample size | 66 | 46 | 45 | 157 |
| Serostable (%, n/N) (Seropositive/Seropositive) | 83.3% (55/66) | 67.4% (31/46) | 64.4% (29/45) | 73.2% (115/157) |
| Seroreverted (%, n/N) (Seropositive/Seronegative) | 16.7% (11/66) | 32.6% (15/46) | 35.6% (16/45) | 26.8% (42/157) |
| Spike OD at enrollment (mean, SD) | 2.73 (0.871) | 1.92 (0.90) | 2.23 (1.17) | 2.35 (1.03) |
| RBD OD at enrollment (mean, SD) | 2.35 (0.91) | 2.29 (0.82) | 2.30 (1.11) | 2.32 (0.94) |
| Days between enrollment and follow up (mean, SD) | 139.4 (14.2) | 123.4 (11.5) | 128.6 (11.8) | 131.6 (14.5) |
| Rate of change Spike OD (OD/100 days) (mean, 95% CI) | -0.11 (-0.32 to 0.10) | -0.03 (-0.319 to 0.259) | -0.15 (-0.34 to 0.04) | -0.10 (-0.23 to 0.03) |
| Rate of change RBD OD (OD/100 days) (mean, 95% CI) | -0.26 (-0.48 to -0.04) | -0.66 (-0.87 to -0.45) | -0.60 (-0.82 to -0.38) | -0.47 (-0.60 to -0.34) |

**Supplementary Table 6: Univariate comparison of serostatus at visit 2 in participants seropositive at visit 1 (n=157, July/October 2020 to December 2021/January 2021)**

| | Seropositive[1] | Seronegative[2] | p-value |
|---|---|---|---|
| Sample size | 115 | 42 | |
| Co-enrolled infants | 0 | 0 | |
| **Demographics** | | | |
| Sex, male (%, n/N) | 36.5% (42/115) | 54.8% (19/42) | **0.0456** |
| Age, years (median, IQR) | 20 (10-39.5) | 11.5 (8-28) | 0.0691 |
| Age group (%, n/N) | | | |
| <10 years | 21.7% (25/115) | 33.3% (14/42) | |
| 10-17 years | 20.9% (24/115) | 30.9% (13/42) | |
| >/=18 years | 57.4% (66/115) | 35.7% (15/42) | |
| Days between enrollment and follow up (mean, SD) | 132.4 (14.9) | 129.3 (13.2) | 0.2287 |
| **Medical factors (% (n/N)** | | | |
| Any comorbidity | 2.6% (3/115) | 0% (0/42) | 0.5641 |
| Pregnancy (any stage) | 1.7% (2/115) | 0% (0/42) | >0.9999 |
| Smoking | 1.7% (2/115) | 4.8% (2/42) | 0.2905 |
| Antimalarial use | 3.5% (4/115) | 2.4% (1/42) | >0.9999 |
| BCG administration | 80.9% (93/115) | 81.0% (34/42) | >0.9999 |
| **Social factors** | | | |
| Works in healthcare | 5.2% (6/115) | 7.1% (3/42) | 0.7017 |
| Household member works in healthcare | 19.1% (22/115) | 19.0% (8/42) | >0.9999 |
| Household size (mean, SD) | 9.1 (6.1) | 7.1 (3.3) | **0.0495** |
| **Symptoms (%, n/N)** | | | |
| No symptoms since visit 1 | 49.1% (68/115) | 54.8% (23/42) | |
| Symptoms since visit 1 | 40.9% (47/115) | 45.2% (19/42) | 0.7155 |
| **Systemic symptoms** | 23.5% (27/115) | 28.6% (12/42) | 0.5354 |
| Fever | 11.3% (13/115) | 9.5% (4/42) | >0.9999 |
| Chills | 4.3% (5/115) | 2.4% (1/42) | >0.9999 |
| Fatigue | 7.0% (8/115) | 9.5% (4/42) | 0.7347 |
| Myalgia | 7.0% (8/115) | 2.4% (1/42) | 0.4462 |
| Headache | 20.9% (24/115) | 21.4% (9/42) | >0.9999 |
| **Respiratory symptoms** | 31.3% (36/79) | 26.2% (11/42) | 0.6941 |
| Sore throat | 5.2% (6/115) | 2.4% (1/42) | 0.6756 |
| Cough | 18.3% (21/115) | 14.3% (6/42) | 0.6397 |
| Rhinorrhea | 23.5% (27/115) | 21.4% (9/42) | 0.8340 |
| Dyspnea | 0% (0/115) | 2.4% (1/42) | 0.2675 |
| Wheezing | 0% (0/115) | 0% (0/42) | >0.9999 |
| Loss of smell/taste | 3.5% (4/115) | 4.8% (2/42) | 0.5743 |
| Other respiratory symptoms | 0% (0/115) | 0% (0/42) | >0.9999 |
| **Gastrointestinal symptoms** | 9.6% (11/115) | 7.1% (3/42) | 0.7610 |
| Nausea/vomiting | 5.2% (6/115) | 2.4% (1/42) | 0.6756 |
| Abdominal pain | 5.2% (6/115) | 7.1% (3/42) | 0.7017 |
| Diarrhea | 0.9% (1/115) | 0% (0/42) | >0.9999 |
| **Symptom severity** | | | |
| Missed work or school | 23.4% (11/47) | 15.8% (3/19) | 0.7407 |
| Sought medical attention | 57.4% (27/47) | 63.2% (12/19) | 0.7849 |
| Hospitalized | 0% (0/47) | 0% (0/19) | >0.9999 |
| Duration of symptoms (any) (mean, SD) | 4.9 (5.4) | 2.8 (1.9) | 0.1870 |
| Symptomatic at Visit 2 | 29.8% (14/47) | 36.8% (7/19) | 0.5748 |

[1]Seropositive refers to participants seropositive at visit 1 and serostable at visit 2
[2]Seronegative refers to participants seropositive at visit 1 and seronegative at visit 2.

**Supplementary Table 7: Adverse events according to serostatus between visit 1 (July/August 2020) and visit 2 (December 2020/January 2021) in individuals co-enrolled in a clinical trial at the Bancoumana site (n=146)**

| | Seronegative[1] | Seropositive[2] | p-value |
|---|---|---|---|
| Sample size | 85 | 61 | |
| **Adverse event (%, n/N)** | | | |
| **Clinical, possibly COVID-19 related** | | | |
| Abdominal pain | 1.2% (1/85) | 1.6% (1/61) | >0.9999 |
| Bronchitis | 3.5% (3/85) | 1.6% (1/61) | 0.6403 |
| Cough | 1.2% (1/85) | 3.3% (2/61) | 0.5714 |
| Chills | 0% (0/85) | 1.6% (1/61) | 0.4218 |
| Decreased appetite | 0% (0/85) | 1.6% (1/61) | 0.4218 |
| Enteritis | 0% (0/85) | 1.6% (1/61) | 0.4178 |
| Gastroenteritis | 2.4% (2/85) | 0% (0/61) | 0.5102 |
| Headache | 15.3% (13/85) | 11.5% (7/61) | 0.6280 |
| Influenza (clinical) | 2.4% (2/85) | 3.3% (2/61) | >0.9999 |
| Nausea | 1.2% (1/85) | 0% (0/61) | >0.9999 |
| Paronychia | 1.2% (1/85) | 0% (0/61) | >0.9999 |
| Pyrexia | 0% (0/85) | 1.6% (1/61) | 0.4296 |
| Rhinitis | 23.5% (20/85) | 34.4% (21/61) | 0.2619 |
| Sinobronchitis | 3.5% (3/85) | 3.3% (2/61) | >0.9999 |
| **Clinical, other** | | | |
| Back pain | 0% (0/85) | 1.6% (1/61) | 0.4218 |
| Conjunctivitis | 9.4% (8/81) | 3.3% (2/61) | 0.1938 |
| Dental caries | 9.4% (8/85) | 0% (0/61) | 0.0209 |
| Dermatosis | 1.2% (1/85) | 0% (0/61) | >0.9999 |
| Dizziness | 1.2% (1/85) | 0% (0/61) | >0.9999 |
| Ear infection | 0% (0/85) | 1.6% (1/61) | 0.4178 |
| Ecchymosis | 0% (0/85) | 1.6% (1/61) | 0.4178 |
| Eye burns | 1.2% (1/85) | 0% (0/61) | >0.9999 |
| Food poisoning | 1.2% (1/85) | 1.6% (1/61) | >0.9999 |
| Gastritis | 2.4% (2/85) | 6.6% (4/61) | 0.2361 |
| Genitourinary tract infection | 2.4% (2/85) | 3.3% (2/61) | >0.9999 |
| Hemorrhoids | 1.2% (1/85) | 0% (0/61) | >0.9999 |
| Hypertension | 1.2% (1/85) | 0% (0/61) | >0.9999 |
| Malaria | 29.4% (25/85) | 32.8% (20/61) | 0.7179 |
| Oropharyngeal pain | 1.2% (1/85) | 0% (0/61) | >0.9999 |
| Pain | 0% (0/85) | 8.2% (5/61) | **0.0139** |
| Strangulated umbilical hernia | 1.2% (1/85) | 0% (0/61) | >0.9999 |
| Tonsillitis | 0% (0/85) | 1.6% (1/61) | 0.4207 |
| Typhoid fever | 2.4% (2/81) | 3.3% (2/61) | >0.9999 |
| Urticaria | 1.2% (1/85) | 0% (0/61) | >0.9999 |
| Wound | 4.7% (8/84) | 8.2% (5/61) | 0.4913 |
| **Laboratory** | | | |
| Alanine aminotransferase increased | 1.2% (1/85) | 0% (0/61) | >0.9999 |
| Blood creatinine increased | 4.7% (4/85) | 1.6% (1/61) | 0.0836 |
| Hemoglobin decreased | 0% (0/85) | 0% (0/61) | >0.9999 |
| Leukopenia | 4.7% (4/85) | 9.8% (6/61) | 0.2020 |
| Neutropenia | 5.9% (5/85) | 11.5% (7/61) | 0.2397 |
| Thrombocytopenia | 0% (0/85) | 3.3% (2/61) | 0.1729 |
| White blood cell count increased | 1.2% (1/85) | 0% (0/61) | >0.9999 |

[1]Seronegative refers to seronegative individuals at visit 1 and visit 2

[2]Seropositive refers to new seropositive individuals at visit 2 (seronegative at visit 1)

**Supplementary Table 8: Grading of commonly reported adverse events in individuals co-enrolled in a clinical trial at the Bancoumana site (n=146)**

| | Seronegative[1] | Seropositive[2] |
|---|---|---|
| Sample size | 85 | 61 |
| **Adverse event (%, n/N)** | | |
| Headache | 15.3% (13/85) | 11.5% (7/61) |
| Grade 1 | 15.4% (2/13) | 14.3% (1/7_) |
| Grade 2 | 84.6% (11/13) | 85.7% (6/7) |
| Grade 3 | 0% (0/13) | 0% (0/7) |
| Rhinitis | 23.5% (20/85) | 34.4% (21/61) |
| Grade 1 | 5.0% (1/20) | 0% (0/21) |
| Grade 2 | 95.0% (19/20) | 100% (21/21) |
| Grade 3 | 0% (0/20) | 0% (0/21) |
| Malaria | 29.4% (25/85) | 32.8% (20/61) |
| Grade 1 | 0% (0/25) | 0% (0/20) |
| Grade 2 | 96.0% (24/25 | 100% (20/20) |
| Grade 3 | 4.0% (1/25) | 0% (0/20) |

[1]Seronegative refers to seronegative individuals at visit 1 and visit 2

[2]Seropositive refers to new seropositive individuals at visit 2 (seronegative at visit 1)

**Supplementary Table 9: Adverse events according to serostatus between visit 1 (July/August 2020) and visit 2 (December 2020/January 2021) in individuals co-enrolled in a clinical trial at the Donéguébougou site (n=1037)**

| | Seronegative[1] | Seropositive[2] | p-value |
|---|---|---|---|
| Sample size | 785 | 252 | |
| **Adverse event (%, n/N)** | | | |
| **Clinical, potentially COVID-19 related** | | | |
| Abdominal pain | 3.1% (24/785) | 3.6% (9/252) | 0.6817 |
| Arthralgia | 0.5% (4/785) | 0% (0/252) | 0.5775 |
| Bronchitis | 2.3% (18/785) | 3.2% (8/252) | 0.4866 |
| Chills | 0.4% (3/785) | 0.4% (1/252) | >0.9999 |
| Cough | 0.4% (3/785) | 0.8% (2/252) | 0.6002 |
| Decreased appetite | 0.8% (6/785) | 1.2% (3/252) | 0.4604 |
| Diarrhoea | 0.1% (1/785) | 0% (0/252) | >0.9999 |
| Gastroenteritis | 1.4% (11/785) | 2.4% (6/252) | 0.2675 |
| Headache | 9.4% (74/785) | 18.3% (46/252) | **0.0003** |
| Myalgia | 0.6% (5/785) | 0% (0/252) | 0.3436 |
| Nasopharyngitis | 1.5% (12/785) | 2.4% (6/252) | 0.4053 |
| Nausea | 0% (0/785) | 0.4% (1/252) | 0.2430 |
| Oropharyngeal pain | 0.4% (3/785) | 0% (0/252) | >0.9999 |
| Pharyngitis | 10.7% (84/785) | 7.1% (18/252) | 0.1138 |
| Pneumonia | 0.3% (2/785) | 0% (0/252) | >0.9999 |
| Pyrexia | 1.9% (15/785) | 3.6% (9/252) | 0.1474 |
| Rhinitis | 25.1% (197/785) | 33.3% (84/252) | **0.0116** |
| Rhinorrhoea | 0.1% (1/785) | 0% (0/252) | >0.9999 |
| Sinobronchitis | 1.5% (12/785) | 0.4% (1/252) | 0.2069 |
| Vomiting | 0.6% (5/785) | 0% (0/252) | 0.3436 |
| **Clinical, other** | | | |
| Abscess | 0.1% (1/785) | 0% (0/252) | >0.9999 |
| Abscess limb | 0.1% (1/785) | 0% (0/252) | >0.9999 |
| Arthropod sting | 0.1% (1/785) | 0.4% (1/252) | 0.4271 |
| Asthenia | 0.6% (5/785) | 0.8% (2/252) | 0.6795 |
| Back pain | 0.1% (1/785) | 0% (0/252) | >0.9999 |
| Chest pain | 0.1% (1/785) | 0% (0/252) | >0.9999 |
| Conjunctivitis | 0.3% (2/785) | 0.4% (1/252) | 0.5666 |
| Dental caries | 1.1% (9/785) | 3.2% (8/252) | **0.0416** |
| Dermatosis | 0.1% (1/785) | 0% (0/252) | >0.9999 |
| Dizziness | 0.8% (6/785) | 1.6% (4/252) | 0.2681 |
| Dysentery | 0.4% (3/785) | 0.8% (2/252) | 0.6002 |
| Dysmenorrhoea | 0% (0/785) | 0.4% (1/252) | 0.2430 |
| Ear infection | 0.1% (1/785) | 0% (0/252) | >0.9999 |
| Epistaxis | 0.1% (1/785) | 0% (0/252) | >0.9999 |
| Food poisoning | 0.1% (1/785) | 0% (0/252) | >0.9999 |
| Fungal skin infection | 0.1% (1/785) | 0% (0/252) | >0.9999 |
| Furuncle | 0.1% (1/785) | 0.4% (1/252) | 0.4271 |
| Gastritis | 0.1% (1/785) | 1.2% (3/252) | **0.0466** |
| Genital infection | 0.1% (1/785) | 0% (0/252) | >0.9999 |
| Gingivitis | 0.1% (1/785) | 0% (0/252) | >0.9999 |
| Hordeolum | 0.1% (1/785) | 0% (0/252) | >0.9999 |
| Hypertension | 0.1% (1/785) | 0% (0/252) | >0.9999 |
| Infection parasitic | 0.1% (1/785) | 0% (0/252) | >0.9999 |
| Injection site pain | 3.8% (30/785) | 4.8% (12/252) | 0.5812 |
| Ligament sprain | 0.1% (1/785) | 0% (0/252) | >0.9999 |
| Limb injury | 0.4% (3/785) | 0.4% (1/252) | >0.9999 |

| | | | |
|---|---|---|---|
| Malaria | 41.1% (323/785) | 41.7% (105/252) | 0.8834 |
| Mastitis | 0.1% (1/785) | 0.4% (1/252) | 0.4271 |
| Otitis externa | 0.3% (2/785) | 0.4% (1/252) | 0.5666 |
| Otitis media | 0.3% (2/785) | 0.4% (1/252) | 0.5666 |
| Pain | 0.3% (2/785) | 0% (0/252) | >0.9999 |
| Pruritus | 0% (0/785) | 0.4% (1/252) | 0.2430 |
| Sciatica | 0.3% (2/785) | 0% (0/252) | >0.9999 |
| Snake bite | 0.1% (1/785) | 0% (0/252) | >0.9999 |
| Tachycardia | 0.3% (2/785) | 0% (0/252) | >0.9999 |
| Thermal burn | 0.1% (1/785) | 0% (0/252) | >0.9999 |
| Tonsillitis | 0.3% (2/785) | 0.4% (1/252) | 0.5666 |
| Urinary tract infection | 0.5% (4/785) | 0.8% (2/252) | 0.6369 |
| Urticaria | 0.3% (2/785) | 0.8% (2/252) | 0.2498 |
| Wound | 1.3% (10/785) | 1.2% (3/252) | >0.9999 |
| Wound infection | 0.4% (3/785) | 0.8% (2/252) | 0.6002 |
| **Laboratory** | | | |
| Alanine aminotransferase increased | 0.8% (6/785) | 1.6% (4/252) | >0.9999 |
| Blood creatinine increased | 0.8% (6/785) | 1.2% (3/252) | 0.4604 |
| Hemoglobin decreased | 0.3% (2/785) | 1.2% (3/252) | 0.0958 |
| Leukocytosis | 0.5% (4/785) | 0.8% (2/252) | 0.6369 |
| Leukopenia | 5.6% (44/785) | 7.1% (18/252) | 0.3624 |
| Neutropenia | 6.8% (53/785) | 7.9% (20/252) | 0.5712 |
| Thrombocytopenia | 0.6% (5/785) | 1.6% (4/252) | 0.2327 |

[1]Seronegative refers to seronegative individuals at visit 1 and visit 2

[2]Seropositive refers to new seropositive individuals at visit 2 (seronegative at visit 1)

**Supplementary Table 10: Grading of commonly reported adverse events in individuals co-enrolled in a clinical trial at the Donéguébougou site (n=1037)**

| | Seronegative[1] | Seropositive[2] |
|---|---|---|
| Sample size | 85 | 61 |
| **Adverse event (%, n/N)** | | |
| Headache | 9.4% (74/785) | 18.3% (46/252) |
| Grade 1 | 98.6% (73/74) | 97.8% (45/46) |
| Grade 2 | 1.4% (1/74) | 2.2% (1/46) |
| Grade 3 | 0% (0/74) | 0% (0/46) |
| Rhinitis | 25.1% (197/785) | 33.3% (84/252) |
| Grade 1 | 91.4% (180/197) | 90.5% (76/84) |
| Grade 2 | 7.1% (14/197) | 9.5% (8/84) |
| Grade 3 | 1.5% (3/197) | 0% (0/84) |
| Malaria | 41.1% (323/785) | 41.7% (105/252) |
| Grade 1 | 77.1% (249/323) | 78.1% (82/105) |
| Grade 2 | 17.6% (57/323) | 17.1% (18/105) |
| Grade 3 | 5.3% (17/323) | 4.8% (5/105) |

[1]Seronegative refers to seronegative individuals at visit 1 and visit 2

[2]Seropositive refers to new seropositive individuals at visit 2 (seronegative at visit 1)
